# Infant Sero-protection: Comparing Antiretroviral therapy, Maternal and Infant Factors Influence on Infant HIV acquisition in Uganda: A six-year Real-World Perspective

**DOI:** 10.1101/2025.06.20.25329979

**Authors:** Collins Ankunda, Jude Emunyu, Brendah Kyomuhangi, Sharon Namasambi, Conrad Sserunjogi, Iving Mumbere, Jane Nakaweesi

## Abstract

**Background:** Prevention of mother-to-child transmission of HIV (PMTCT) remains a critical pillar in reducing new paediatric HIV infections. While Antiretroviral Therapy (ART) scale-up has significantly reduced transmission, differential effectiveness of ART regimens, maternal and infant factors within PMTCT programs remain underexplored. This study examined the comparative influence of ART, maternal and infant factors on the risk of infant HIV seroconversion.

**Methods:** We retrospectively reviewed records of 962 HIV-exposed infants (918 mother-infant pairs and 44 without maternal data) from the Mildmay Uganda PMTCT clinic. Infant HIV serostatus was analyzed in relation to ART, maternal, and infant characteristics. Descriptive statistics summarized the data; associations were tested using Chi-square and Fisher’s exact tests. Bivariate and multivariable logistic regression identified predictors. The maternal and infant models were compared using log-likelihood, Akaike Information Criterion (AIC), and Bayesian Information Criterion (BIC) statistics.

**Results:** Overall HIV seropositivity among mother-infant pairs was 1.96% (18/918). When disaggregated by maternal ART regimen, the lowest seropositivity occurred with dolutegravir (DTG)-based regimens (1.4%) and the highest with protease inhibitor (PI)-based regimens (5.88%). Differences across regimens were not statistically significant (p=0.113). Among maternal factors, age above 35 years (p=0.000) and unsuppressed viral load (p=0.025) were significantly associated with higher seropositivity. Evaluation of Infant-related factors showed that no nevirapine prophylaxis (aOR=43.74; 95% CI: 17.88–106.96; p=0.000), no cotrimoxazole prophylaxis (aOR=11.45; 95% CI: 2.54–51.66; p=0.002), mixed feeding (aOR=26.87; 95% CI: 5.82–124.10; p=0.000) and no breastfeeding (aOR=86.94; 95% CI: 17.40–434.56; p=0.000) were independently associated with MTCT. The maternal model (AIC = 160.09; BIC = 207.19) showed a better fit over the infant model (AIC = 185.91; BIC = 219.99). However, only the infant model identified statistically significant predictors of HIV seropositivity.

**Conclusions:** Our findings encourage maintaining similar regimens during pregnancy and breastfeeding unless clinically indicated. While maternal ART regimen and viral load suppression are key in reducing MTCT, infant-related factors such as prophylaxis and feeding practices more strongly predict infant HIV risk. Integrated PMTCT approaches must ensure sustained scale up of ART, maternal viral suppression, promote exclusive breastfeeding, and guarantee infant prophylaxis to achieve the goal of eliminating paediatric HIV.

## Background

Globally, Mother-to-child transmission (MTCT) remains a major source of paediatric HIV, particularly in sub-Saharan Africa[1], where the burden of HIV is high, but ART scale-up among pregnant women living with HIV (PWLH) has significantly reduced transmission risk during pregnancy, delivery, and breastfeeding. Without intervention, MTCT risk ranges from 15% to 45%[1], but with effective ART and comprehensive PMTCT measures, it can be reduced to under 5%, or even below 2% in some settings[1–3].

Several maternal factors influence MTCT risk during and after pregnancy, including ART use, viral load, overall health, breastfeeding practices, and sexually transmitted infections (STIs) like syphilis[4,5]. High maternal viral load at delivery is a key transmission driver[6]. Effective ART regimens that rapidly and sustainably suppress viral load are essential[7,8], highlighting the importance of timely initiation, adherence, and robust maternal healthcare to reduce MTCT risk, favouring drugs that achieve rapid and sustained viral suppression[9,10]. Dolutegravir (DTG)-based regimens have become the preferred first-line treatment for PWLH due to their superior efficacy in rapidly suppressing viral load compared to previous regimens, such as efavirenz (EFV)-based combinations[2,11,12]. The early and sustained suppression of maternal viral load during pregnancy significantly lowers the risk of intrauterine and peripartum transmission of HIV[10].

Postpartum MTCT risk is strongly influenced by breastfeeding practices and maternal viral suppression[13,14]. Infant feeding practices are pivotal; the World Health Organization recommends exclusive breastfeeding for the first six months, followed by complementary feeding while continuing breastfeeding up to one year or beyond, depending on maternal viral suppression[15]. Infant antiretroviral prophylaxis, such as nevirapine, is tailored to exposure risk[2]. Sustained maternal viral suppression throughout breastfeeding remains critical for minimizing postnatal HIV transmission in exposed infants[10].

While the effectiveness of PMTCT interventions has been well documented, most studies have focused on the overall impact of ART and infant prophylaxis without disaggregating the contributions of antepartum and postpartum interventions. Limited research has systematically compared the relative contributions of interventions during the antepartum period and the postpartum period to HIV seroconversion among infants. The study compared the impact of antepartum and postpartum factors that influence infant HIV seroconversion among PWLH on different ART regimens. We examined the influence of these factors across the maternal-infant care continuum to inform strategies for optimizing PMTCT outcomes and achieving the goal of eliminating paediatric HIV infections in high HIV burden setting.

## Methods and materials

### Study Design

This cohort study retrospectively reviewed medical records of PWLH enrolled in the PMTCT clinic at the Mildmay Uganda Hospital (MUgH) from 2018 to 2023.

### Study site and population

This study was conducted at MUgH, a peri-urban facility in Wakiso District with 50-bed capacity, providing integrated TB-HIV care and ART to nearly 15,000 active HIV patients[16]. The study population included pregnant women, breastfeeding mothers and HIV-exposed infants with final HIV test results in accordance with Ministry of Health guidelines[2]. Mothers who gave birth more than once during the six-year study period were treated as independent observations.

### Data Collection and Management

Data collection and participant recruitment were conducted over a three-month period (1st July to 30th September 2024). Data on participants’ socio-demographic characteristics and medical history were extracted from PMTCT registers, patient cards, and Electronic Medical Records (EMR). Data entry was performed using Microsoft Excel, with regular accuracy checks against hard copy records to ensure data integrity.

A flowchart illustrates the data collection process.

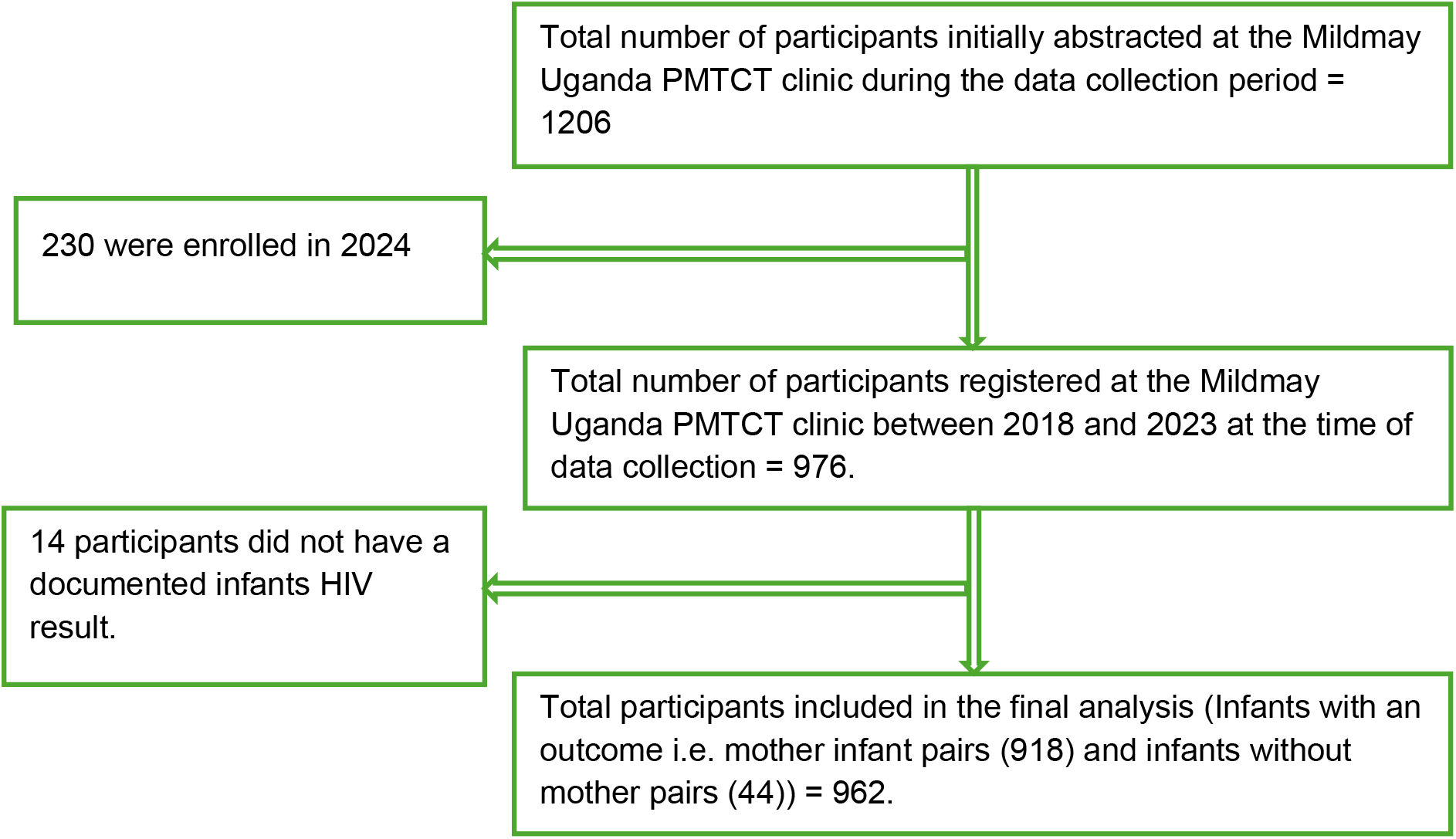

### Statistical Analysis Plan

Data analysis was conducted using Stata 16. The primary outcome was infant HIV seroconversion, treated as a binary variable(seroconverted-Yes/No). Influence of ART regimen was assessed using Fisher’s exact (only mother infant pairs were considered). Analysis of maternal and infant factors was done using descriptive statistics using frequencies and percentages, with categorical variables compared via Chi-square or Fisher’s exact tests as appropriate. Bivariate logistic regression identified variables associated with infant HIV seroconversion at p < 0.20 for inclusion in multivariable models. A hierarchical logistic regression approach was then used to evaluate the effects of maternal and infant factors. Crude (OR) and Adjusted odds ratios (aOR) with 95% confidence intervals (CI) were reported. Statistical significance was set at p < 0.05. Comparison between maternal and infant factors was done. Model 1 included maternal (antepartum) factors while Model 2 included infant (postpartum) factors. Model fit was assessed using Akaike Information Criterion (AIC) and Bayesian Information Criterion (BIC) with lower values considered a better model fit.

### Ethical Considerations

Ethical approval was obtained from the Mildmay Uganda Research Ethics Committee (#REC REF 0201-2024) and the Uganda National Council for Science and Technology (HS3873ES). A waiver of informed consent was granted by Mildmay Uganda Research Ethics Committee for the use of de-identified retrospective data. Administrative clearance was sought from MUgH administration to access participant data.

## Results

### Influence of PMTCT ART Regimen on infant HIV seropositivity

Among 918 HIV-exposed infants with mother pairs, 18 (1.96%, 95% CI: 1.15–3.06) were confirmed HIV seropositive. When stratified by maternal ART regimen, seropositivity was 1.40% (8/571; 95% CI: 0.61–2.74) for infants whose mothers were on DTG-based regimens, 2.96% (8/270; 95% CI: 1.29–5.75) for those on Efavirenz(EFV)-based regimens, 0% (0/43;95% CI: 0.00–8.22) for Nevirapine(NVP)-based regimens, and 5.88% (2/34; 95% CI: 0.72– 19.68) for Protease inhibitor(PI)–based regimens. The difference in seropositivity rates across these ART regimens was not statistically significant (p = 0.113) as shown in table 1 below.

**Table 1:**
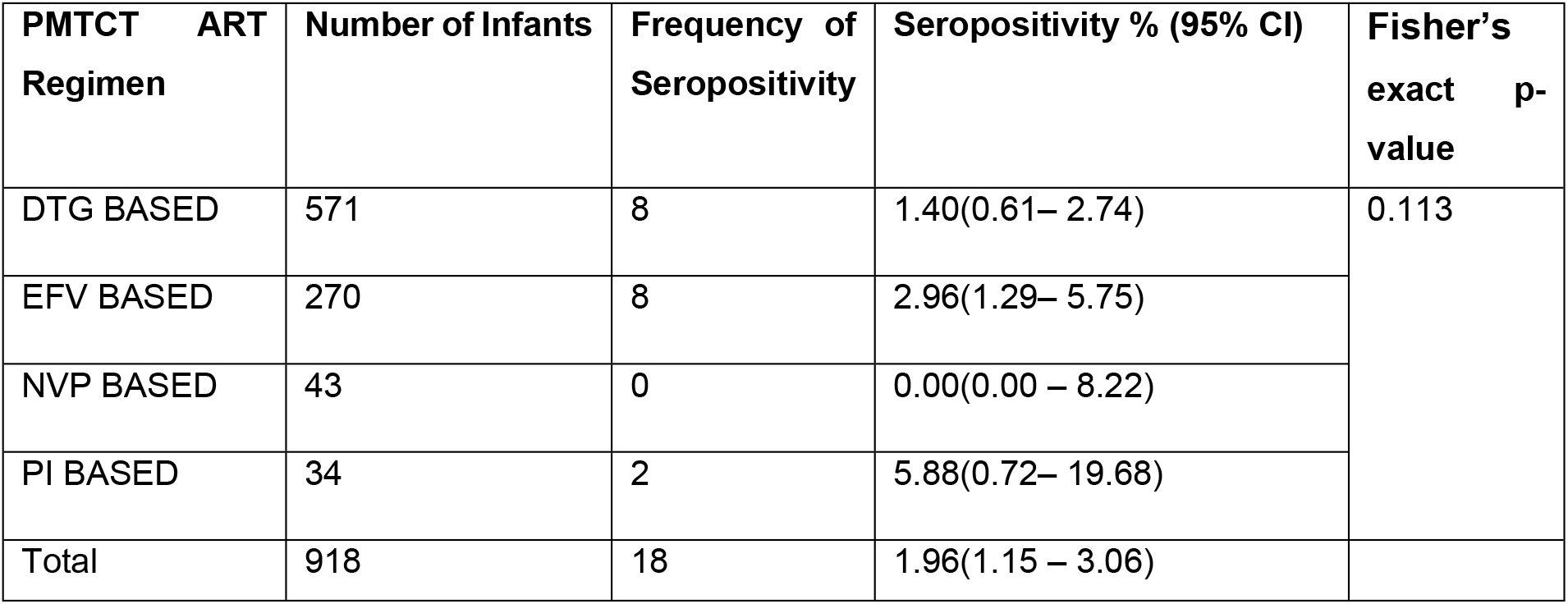
Proportion of HIV seropositivity by PMTCT ART Regimen Category.

| PMTCT ART Regimen | Number of Infants | Frequency of Seropositivity | Seropositivity % (95% CI) | Fisher's exact p-value |
| --- | --- | --- | --- | --- |
| DTG BASED | 571 | 8 | 1.40(0.61– 2.74) | 0.113 |
| EFV BASED | 270 | 8 | 2.96(1.29– 5.75) |  |
| NVP BASED | 43 | 0 | 0.00(0.00 – 8.22) |  |
| PI BASED | 34 | 2 | 5.88(0.72– 19.68) |  |
| Total | 918 | 18 | 1.96(1.15 – 3.06) |  |

### Distribution of Infant HIV Seropositivity by Maternal and Infant Characteristics

Table 2 shows that among the 918 mother-infant pairs, key maternal factors significantly associated with infant HIV seropositivity included maternal age (p=0.000), with the highest positivity among women over 35 years (5.8%), and PWLH with a non suppressed viral load (6.7% vs. 1.6%, p=0.025). When all the 962 infants were assessed, all infant-related factors were significantly associated with seroconversion. Infants not receiving nevirapine prophylaxis had a markedly higher seropositivity rate (53.7% vs. 2.3%, p=0.000). Similarly, those not on cotrimoxazole had a 50% positivity rate compared to 4.1% among those who received it (p=0.000). Feeding practices also impacted outcomes: exclusive breastfeeding had the lowest seropositivity (0.4%), while mixed and no breastfeeding had significantly higher rates (9.1% and 25.0%, respectively; p=0.000).

**Table 2.** Distribution of Infant HIV Seropositivity by Maternal and Infant Characteristics.

| Group Characteristics | Variable | Categories | Negative | Positive | P-value** |
| --- | --- | --- | --- | --- | --- |
| Maternal Characteristics | Age (Years) | 15-24 | 114(100.0) | 0(0.0) | **0.000** |
|  |  | 25-34 | 510(99.8) | 1(0.2) |  |
|  |  | >35 | 276(94.2) | 17(5.8) |  |
|  | WHO stage | Stage 1 | 847(97.9) | 18(2.1) | 1.000 |
|  |  | Stage 2 | 19(100.0) | 0(0.0) |  |
|  |  | Stage 3 | 9(100.0) | 0(0.0) |  |
|  |  | Stage 4 | - | - |  |
|  | Adherence | Good | 889(98.1) | 17(1.9) | 0.181 |
|  |  | Fair | 2(100.0) | 0(0.0) |  |
|  |  | Poor | 7(87.5) | 1(12.5) |  |
|  | Start Regimen | DTG | 119(98.4) | 2(1.6) | 0.550 |
|  |  | EFV | 605(98.2) | 11(1.8) |  |
|  |  | NVP | 143(98.0) | 3(2) |  |
|  |  | PI | 19(95.0) | 1(5.0) |  |
|  | Drug line | First line | 819(98.1) | 16(1.9) | 0.678 |
|  |  | Second line | 80(97.6) | 2(2.4) |  |
|  |  | Third line | 1(100.0) | 0(0.0) |  |
|  | Duration on ART (years) | >1 | 26(92.9) | 2(7.1) | 0.114 |
|  |  | 1-2 | 73(97.3) | 2(2.7) |  |
|  |  | >2-5 | 243(97.6) | 6(2.4) |  |
|  |  | >5 | 558(98.6) | 8(1.4) |  |
|  | History of regimen change | No | 109(98.2) | 2(1.8) | 1.000 |
|  |  | Yes | 791(98.0) | 16(2.0) |  |
|  | ART experience | Experienced | 872(98.2) | 16(1.8) | 0.114 |
|  |  | Naive | 28(93.3) | 2(6.7) |  |
|  | Viral load suppression | Suppressed | 844(98.4) | 14(1.6) | **0.025** |
|  |  | Unsuppressed | 56(93.3) | 4(6.7) |  |
|  | Marital status | Married | 433(97.7) | 10(2.3) | 0.854 |
|  |  | Single | 303(98.4) | 5(1.6) |  |
|  |  | Separated/Divorced/Widowed | 79(97.5) | 2(2.5) |  |
|  |  | Unknown | 85(98.8) | 1(1.2) |  |
|  | Mode of delivery | Caesarean section | 242(98.4) | 4(1.6) | 1.000 |
|  |  | SVD | 637(98.0) | 13(2.0) |  |
|  | Place of delivery | Health facility | 896(98.0) | 18(2.0) | 1.000 |
|  |  | Home/ Unknown | 22(100.0) | 0(0.0) |  |
|  | History of ART transition* | 0 | 448(98.9) | 5(1.1) | 0.112 |
|  |  | 1 | 19(100.0) | 0(0.0) |  |
|  |  | 2 | 27(96.4) | 1(3.6) |  |
|  |  | 3 | 3(100.0) | 0(0.0) |  |
|  |  | 4 | 107(98.2) | 2(1.8) |  |
|  |  | 5 | 237(96.7) | 8(3.3) |  |
|  |  | 6 | 5(83.3) | 1(16.7) |  |
|  |  | 7 | 40(100.0) | 0(0.0) |  |
| Infant Characteristics | Received NVP syrup | Yes | 900(97.7) | 21(2.3) | 0.000 |
|  |  | No/Not sure | 19(46.3) | 22(53.7) |  |
|  | Received Cotrimoxazole prophylaxis | Yes | 915(95.9) | 39(100.0) | 0.000 |
|  |  | No/not sure | 4(50.0) | 4(50.0) |  |
|  | Infant Feeding option at the time of first test | Exclusive Breast feeding | 806(99.6) | 3(0.4) | 0.000 |
|  |  | Mixed feeding | 40(90.9) | 4(9.1) |  |
|  |  | Not breast feeding | 12(75.0) | 4(25.0) |  |
|  |  | Replacement feeding | 35(97.2) | 1(2.8) |  |
|  |  | Unknown | 26(45.6) | 31(54.4) |  |

\*The study categorized antiretroviral therapy (ART) exposure based on maternal regimen transitions during pregnancy and breastfeeding. These included: transition from other regimens to DTG-based (0), EFV-based (1), PI-based (2), or NVP-based (3) regimens, as well as cases where PWLH remained on the same regimen throughout; DTG-based (4), EFV-based (5), PI-based (6), or NVP-based (7).
\*\*P-values were derived using Chi-square tests, or Fisher's exact tests when cell counts were < 5.

### Bivariate and Multivariate Analysis of Maternal and Infant Factors Associated with MTCT of HIV

In bivariate analysis, maternal age 25–34 years was associated with significantly lower odds of MTCT compared to >35 years (OR=0.03; 95% CI: 0.00–0.24; p=0.001). Remaining on PI regimen increased MTCT odds (OR=17.92; 95% CI: 1.76–182.51; p=0.015). Among infants, not receiving nevirapine syrup (OR=49.62; 95% CI: 23.42–105.16; p<0.001), not receiving cotrimoxazole prophylaxis (OR=23.46; 95% CI: 5.66–97.30; p<0.001), and feeding practices; mixed feeding (OR=26.87; 95% CI: 5.82–124.11; p<0.001), no breastfeeding (OR=89.56; 95% CI: 18.05–444.37; p<0.001), and unknown feeding option (OR=320.33; 95% CI: 91.98– 1115.58; p<0.001) were significantly associated with MTCT. In the multivariate model, mixed feeding (aOR=26.87; 95% CI: 5.82–124.10; p<0.001), no breastfeeding (aOR=86.94; 95% CI: 17.40–434.56; p<0.001), and unknown feeding (aOR=275.00; 95% CI: 66.04–1145.72; p<0.001) remained independently associated with MTCT (Table 3).

**Table 3:** Bivariate and Multivariate Analysis of Maternal and Infant Factors Associated with MTCT of HIV.

| Models | Variable | Categories | OR (95% CI) | p-value | aOR (95% CI) | P-Value |
| --- | --- | --- | --- | --- | --- | --- |
| **Maternal predictors** | PMTCT ART Regimen | DTG | Ref |  | Ref |  |
|  |  | EFV | 2.19(0.81-5.91) | 0.120 | 2.02(0.52-7.89) | 0.309 |
|  |  | PI | 4.32(0.88-21.18) | **0.071** | 15.93(0.28-890.60) | 0.178 |
|  |  | NVP | 1 | - |  |  |
|  | Age (Years) | 15-24 | 1 | - |  |  |
|  |  | 25-34 | 0.03(0.00-0.24) | **0.001** | - | - |
|  |  | >35 | Ref |  |  |  |
|  | Adherence | Good | Ref |  |  |  |
|  |  | Fair | 1 | - |  |  |
|  |  | Poor | 7.47(0.87-64.10) | 0.067 | - | - |
|  | Regimen at ART initiation | DTG | Ref |  |  |  |
|  |  | EFV | 1.08(0.24-4.94) | 0.919 |  |  |
|  |  | PI | 3.13(0.27-36.24) | 0.361 |  |  |
|  |  | NVP | 1.25(0.21-7.59) | 0.810 |  |  |
|  | Drug line during PTCMT | First line | Ref |  |  |  |
|  |  | Second line | 1.31(0.30-5.80) | 0.721 |  |  |
|  |  | Third line | 1 | - |  |  |
|  | Duration on ART (years) | >1 | 3.12(0.60-16.23) | 0.177 | 9.53(0.93-97.95) | 0.058 |
|  |  | 1-2 | 1.11(0.22-5.62) | 0.900 | 0.83(0.07-10.32) | 0.887 |
|  |  | >2-5 | Ref |  | Ref |  |
|  |  | >5 | 0.58(0.20-1.69) | 0.319 | 0.41(0.98-1.71) | 0.221 |
|  | History of regimen change | No | Ref |  |  |  |
|  |  | Yes | 1.09(0.25-4.82) | 0.906 |  |  |
|  | ART experience | Experienced | Ref |  |  |  |
|  |  | Naive | 3.89(0.85-17.75) | 0.079 | - |  |
|  | Viral load suppression | Suppressed | Ref |  | Ref |  |
|  |  | Unsuppressed | 3.89(0.85-17.75) | 0.079 | 2.91(0.43-19.57) | 0.272 |
|  | Marital status | Married | Ref |  |  |  |
|  |  | Single | 0.71(0.24-2.11) | 0.543 |  |  |
|  |  | Separated/Divorced /Widowed | 1.10(0.24-5.10) | 0.907 |  |  |
|  |  | Unknown | 0.51(0.06-4.03) | 0.523 |  |  |
|  | Mode of delivery | Caesarean section | Ref |  |  |  |
|  |  | SVD | 1.23(0.40-3.82) | 0.715 |  |  |
|  | ART history of transition | 0 | Ref |  | Ref |  |
|  |  | 1 | 1 | - | 1 | - |
|  |  | 2 | 3.32(0.37-29.41) | 0.281 | 0.33(0.00-28.94) | 0.628 |
|  |  | 3 | 1 | - | 1 | - |
|  |  | 4 | 1.67(0.32-8.75) | 0.541 | 0.26(0.02-3.48) | 0.309 |
|  |  | 5 | 3.02(0.98-9.35) | 0.055 | 1 | - |
|  |  | 6 | 17.92(1.76-182.51) | 0.015 | 1 | - |
|  |  | 7 | 1 | - | 1 | - |
| **Infant predictors** | Received NVP syrup | Yes | Ref |  | Ref |  |
|  |  | No/Not sure | 49.62(23.42-105.16) | **0.000** | 1.25(0.43-3.69) | 0.682 |
|  | Received Cotrimoxazole prophylaxis | Yes | Ref |  | Ref |  |
|  |  | No/not sure | 23.46(5.66-97.30) | **0.000** | 1.02(0.20-5.22) | 0.983 |
|  | Infant Feeding option at the time of first test | Exclusive Breast feeding | Ref |  | Ref |  |
|  |  | Mixed feeding | 26.87(5.82-124.11) | **0.000** | 26.87(5.82-124.10) | **0.000** |
|  |  | Not breast feeding | 89.56(18.05-444.37) | **0.000** | 86.94(17.40-434.56) | **0.000** |
|  |  | Replacement feeding | 7.68(0.78-75.68) | 0.081 | 7.62(0.77-75.16) | 0.082 |
|  |  | Unknown | 320.33(91.98-1115.58) | **0.000** | 275(66.04-1145.72) | **0.000** |

### Comparison of Maternal and Infant Predictors of MTCT of HIV

Assessment for the best fit model to predict MTCT of HIV was done. The maternal model, with 821 observations, had a log-likelihood of -70.04, 10 degrees of freedom, AIC of 160.09, and BIC of 207.19. The infant model included 962 observations, log-likelihood of -85.95, 7 degrees of freedom, AIC of 185.91, and BIC of 219.99. The maternal model showed a lower AIC and BIC indicating better model fitness as shown in table 4 below.

**Table 4:**
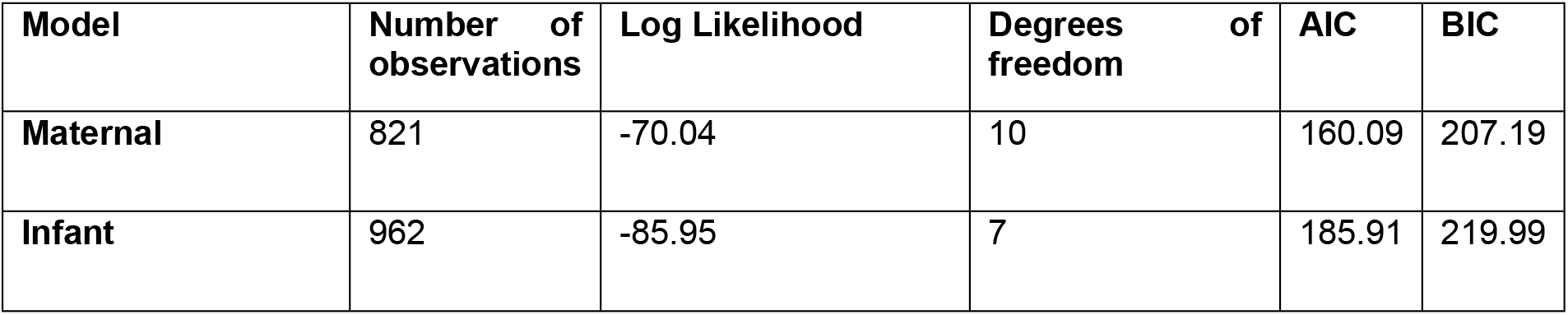
Model fit statistics for maternal and infant predictors of MTCT of HIV.

## Discussion

This study comprehensively examined the relative contributions of maternal and infant factors in influencing mother-to-child transmission (MTCT) of HIV within the Ugandan context. With an overall infant seropositivity rate of 1.96% across maternal ART regimens, our findings highlight the significant progress achieved through widespread PMTCT intervention[1–3] while emphasising ongoing challenges in HIV exposed infant care. Additionally, we examined HIV seropositivity across maternal ART regimens and observed key clinical trends. No variation in seropositivity was observed DTG, EFV, PI and NVP based regimens. These findings suggest that DTG-, EFV-, and NVP-based regimens offer comparable protection against mother-to-child HIV transmission[17]. However, the small sample sizes in the PI and NVP groups limit definitive conclusions.

Evaluation of maternal factors revealed that being aged 25 to 34 years was significantly associated with transmitting HIV compared to having over 35 years, suggesting a higher MTCT risk with increasing maternal age. Similarly, a study done in India reported that women aged 30 years and above had a sevenfold increased risk of MTCT [19]. While previous literature often associates older age with better antiretroviral therapy (ART) outcomes due to presumed greater maturity, awareness, and adherence [17,18], this trend was not observed in our cohort. Age specific interventions may play a role in enhancing PMTCT outcomes. Further more, our study showed that unsuppressed maternal viral load was associated with MTCT risk, consistent with the established link between maternal viremia and infant infection[2,18,19]. This emphasises viral load suppression as a cornerstone of PMTCT success highlighting the need for routine viral load monitoring among pregnant and breast-feeding women for better exposed HIV infant outcomes[2].

All Infant-related factors showed statistically significant associations with HIV seroconversion, emphasizing their critical role in prevention of postnatal HIV transmission. Infants who did not receive nevirapine prophylaxis had an increased risk of HIV seropositivity, affirming nevirapine’s protective efficacy in blocking viral replication during the vulnerable early postnatal period[20]. Similarly, absence of cotrimoxazole prophylaxis, an intervention primarily targeting opportunistic infections, was linked to markedly higher seropositivity. Although cotrimoxazole’s direct role in preventing MTCT is less defined[21,22], its use can be a proxy indicator of health system engagement, quality infant care and maternal adherence to prescribed instructions, which may indirectly reduce the risk of HIV transmission. Notably, infant feeding practices emerged as the most decisive infant factor, with exclusive breastfeeding associated with the lowest HIV seropositivity. In contrast, mixed feeding and no breastfeeding had significantly higher transmission rates. These findings are consistent with biological mechanisms: mixed feeding can damage the infant gut mucosa, facilitating viral entry, while exclusive breastfeeding maintains gut integrity and transmits protective maternal antibodies[23]. The heightened risk in infants not breastfed may relate to lack of immune protection and replacement feeding challenges in resource-limited settings[2,23]. The high odds to seropositivity among infants with unknown feeding practices may highlight data gaps and possible non-adherence to recommended feeding guidelines.

The study compared two logistic regression models to assess the better predictor of MTCT. The maternal model showed a better overall fit based on AIC and BIC values. This suggests that maternal factors may be more informative in predicting MTCT than infant characteristics. This is supported by the emphasis on strengthening maternal care in PMTCT programs[2]. However, only the infant model contained statistically significant predictors of MTCT. This suggests that while maternal factors may offer a better platform for evaluation for risk of HIV transmission, infant care practices exert a stronger direct influence on seropositivity. This is explained by fact that maternal ART and viral suppression predominantly mitigate intrauterine and intrapartum transmission risks, but postnatal factors like, infant prophylaxis and feeding, heavily influence seropositivity during the period of continued exposure[24]. This calls for dual focus in PMTCT programs: maintaining maternal viral suppression through early ART initiation, adherence, and monitoring, alongside ensuring robust infant prophylaxis and counselling on safe feeding practices. Enhanced health education to promote exclusive breastfeeding and discourage mixed feeding must remain a priority.

### Implications for Policy and Practice

Our findings demonstrate the importance of scaling up all ART regimens except the PI-based in PMTCT programs. Policy should avoid unnecessary regimen switch or transition among pregnant and breast-feeding women unless clinically indicated.

Second, special attention is warranted for older mothers and those with unsuppressed viral loads, who represent a higher-risk group. Interventions such as targeted adherence counselling, peer to peer psychosocial support, viral load monitoring, and regimen optimization could reduce their transmission risk.

Importantly, during the post partum period, infant prophylaxis coverage and exclusive breastfeeding promotion are key actionable areas. Also, PMTCT programs should ensure timely initiation of nevirapine and cotrimoxazole for all HIV-exposed infants.

### Strengths and Limitations

This study’s strengths include a large sample size and clinic-based data collected over six years, offering a comprehensive evaluation of both maternal and infant factors within a real-world clinical context. This enhances the generalizability of the findings. However, some limitations must be acknowledged. The inherent limitations of a retrospective design, including the inability to infer causality and potential bias from incomplete data, may have affected the accuracy of some point estimates. Additionally, small numbers within certain ART regimen subgroups limited statistical power, while self-reported infant feeding practices may be affected by recall or social desirability bias. Despite these constraints, the study provides important insights into the relative contributions of maternal and infant factors to MTCT and identifies key areas for strengthening the PMTCT program.

## Conclusion

In summary, while maternal factors such as ART regimen and viral load suppression remain foundational in preventing MTCT, infant factors particularly prophylaxis and feeding practices play a more decisive role in infant HIV risk. These findings emphasize the need for integrated approaches that concurrently optimize maternal treatment and infant care to achieve the goal of eliminating paediatric HIV.

Targeted strategies promoting exclusive breastfeeding, ensuring prophylaxis coverage, and supporting maternal viral suppression, especially among older women, are essential. Strengthening health systems to improve facility-based delivery and postnatal follow-up will further augment PMTCT programs. Continued research and programmatic innovation remain critical in sustaining and accelerating progress toward an HIV-free generation in Uganda and beyond.

## Data Availability

The data sets for this study are available upon reasonable request from the corresponding author.

## Acknowledgments

The authors extend their gratitude to the study participants, hospital administrators, staff, and healthcare workers. Their contributions at various levels were instrumental in making this study a success.

